# Recovery following femoral periprosthetic fracture: a qualitative study of patient experience and socio-economic influences during acute hospitalisation

**DOI:** 10.64898/2026.01.19.25343124

**Authors:** Lewis Navarro

## Abstract

**Background:** Femoral periprosthetic fractures (PPFs) are a significant complication following hip and knee arthroplasty, with an incidence of approximately 5.5–18%. These injuries require complex surgical management and are associated with persistent pain, impaired mobility, prolonged hospital stays, and poorer functional outcomes. Socio-economic factors are known to influence recovery, yet patient perspectives following PPFs remain underexplored.

**Aim:** To explore patients’ experiences of recovery following femoral periprosthetic fracture during hospital admission, with particular attention to the influence of socio-economic factors.

**Methods:** A qualitative study design was employed using semi-structured interviews with patients recovering from femoral PPFs during inpatient admission. Participants (n=9) were recruited from a major trauma hospital in the North West of England and categorised into socio-economic groups using the English Indices of Deprivation. Data were analysed using Braun and Clarke’s thematic analysis framework, supported by NVivo® software, with both inductive and deductive coding approaches.

**Results:** Seven core themes were identified across two overarching categories: Recovery Experience and Socio-economic Influences. Recovery experience included themes of persistent pain, restricted mobility, emotional distress, and a strong desire to regain independence characterised recovery experiences. Whereas, socio-economic factors influenced access to resources, communication with healthcare professionals, and reliance on family and social support networks. Participants from more socio-economically deprived backgrounds reported greater frustration related to delays, perceived resource limitations, and unmet recovery needs.

**Conclusions:** Recovery following femoral PPF is shaped by an interaction of physical, psychological, and socio-economic factors during hospitalisation. These findings highlight the need for integrated and individualised care pathways that explicitly address socio-economic inequalities. Improved communication, tailored rehabilitation planning, and enhanced support mechanisms may contribute to more equitable and meaningful recovery outcomes for this vulnerable patient population.

**Lay Summary:** *Study Title:* Recovery following femoral periprosthetic fracture: a qualitative study of patient experience and socio-economic influences during acute hospitalisation

*Who carried out the research?:* This study was conducted by a researcher based at a major trauma hospital in the North West of England. The study was part of a postgraduate academic project. There were no commercial sponsors or competing interests.

*Thank you to study participants:* We would like to sincerely thank the patients who took part in this study. Their time, honesty, and openness helped us better understand what recovery really feels like after a major injury.

*Why was the research needed?:* A femoral periprosthetic fracture is a serious complication that can happen after hip or knee replacement surgery. It often leads to a long recovery, pain, and difficulty returning back to everyday activities such as walking. We know that people from different backgrounds don’t always have the same access to care or support, which can affect how well they recover. However, very little is known about patients’ personal experiences, especially those from more deprived areas. This study aimed to fill that gap.

*What were the main questions studied?:*

1. What is it like for patients recovering from a femoral periprosthetic fracture while in hospital?
2. Do patients from different socio-economic backgrounds experience recovery in different ways?

*Where and when did the study take place?:* The study took place in 2024 at a major NHS trauma hospital in the North West of England. Interviews were conducted with patients while they were still in hospital.

*Who participated in the study?:* Nine adult patients recovering in hospital after a femoral periprosthetic fracture took part. They were grouped according to where they lived, based on national measures of deprivation. This allowed the researcher to explore the effects of social and economic background.

*What happened during the study?:* Participants took part in a one-on-one interview with the researcher. They were asked open questions about their recovery, how they felt physically and emotionally, what support they had, and how they were being treated during their hospital stay. The responses were analysed to find common themes and differences.

*What were the results of the study?:* The study found four key themes grouped under two main areas: the general recovery experience and the impact of socio-economic background.

- Most patients reported **ongoing pain**, **problems with mobility**, and **emotional distress**, such as anxiety or feeling a loss of independence.
- Many said that **regaining independence** — being able to move freely or go outside on their own — was their top recovery goal.
- **Family support** was essential. Those without strong social support, particularly from more deprived backgrounds, often struggled more.
- Patients from lower-income regions reported **more delays**, **worse communication**, and **fewer resources** available to help them recover.

*How has this study helped patients and researchers?:* This study gives a voice to patients recovering from a complex injury and highlights how social and economic inequality can influence health outcomes. It shows the need for:

- Better emotional and psychological access and support during recovery following injury.
- Recovery goals that are personalised and focused on independence.
- More support systems (e.g., community care) for people without family or friend help.
- Clearer, more compassionate communication from healthcare staff particularly when awaiting surgery.

*Details of any further research planned.:* Future studies should involve a larger and more diverse group of patients, follow their recovery over a longer period (including after discharge), and test specific solutions such as improved communication or social support programs whilst in hospital.

*Where can I learn more about this study?:* You can find more information by contacting the research team at Lancashire Teaching Hospitals NHS Foundation Trust, Royal Preston Hospital, Core Therapies Department or by accessing future published versions of this study in academic journals or NHS research summaries.

## Introduction

Femoral Periprosthetic Fractures (PPFs) are a significant complication following joint arthroplasties, such as hip or knee replacements, with an incidence range of 5.5% to 18% which is a reported two and half-fold increase over the past twenty years (Sadoghi et al., 2013; Young et al., 2007; Frenzel et al., 2015). Femoral PPF’s are known to be the third highest cause for revision surgery within two years of joint arthroplasty and the second highest cause for revision thereafter (AOANJRR, 2018). The rising incidences are associated with increasing frequency of joint replacement surgeries in addition to the increase in average life-expectancy (Frenzel et al., 2015). Other observational associations include surging numbers of uncemented femoral fixations and revision total hip replacements (Abdel et al., 2018). With the aging global population combined with increasing contributing surgeries, an ever-larger population is at risk.

Typically, femoral PPF’s require urgent surgical intervention, sometimes with added open reduction internal fixation (ORIF), often involving implant revision or joint replacement; however, non-operative measures are also known to be considered (Abdel et al., 2016; Morgan et al., 2023). Despite advances in femoral PPF classification and surgical techniques, recovery outcomes remain poor even without the added associative risk of complications such as joint dislocation, malunion, infection and further surgery (Holder et al., 2014). Studies report that nearly half of patients experience complications, fail to regain pre-injury mobility, face prolonged hospital stays, and have a significantly higher mortality risk compared to primary hip replacements (Bhattacharyya et al., 2007; Della Rocca et al., 2011; Moreta et al., 2014; Streubel PN. 2013).

There is a significant burden associated with femoral PPFs which extends much further than just the healthcare system and the patient’s functional recovery. A sole biomedical approach discredits the substantial importance in psychosocial factors in recovery from femoral PPFs (Griffiths et al., 2015; Schiller et al., 2015; Schemitsch & Nauth, 2020). Aspects such as pain, dependency, and disability often lead to psychological disorders such as anxiety, depression, and post-traumatic stress disorder (PTSD) and are known to be undervalued and undertreated contributors to chronic pain, long-term disability, poor functional outcome and poor Quality of Life (QoL) at 3 and 24 months following orthopaedic trauma (Demnati et al., 2024; Islam et al., 2022; Schemitsch & Nauth 2020; Sjoerd et al., 2015). Despite this, orthopaedic trauma research in femoral PPFs continues to be extensively focused on surgical, physical and technical components.

Moreover, it is increasingly apparent that socio-economic factors also play a critical role in shaping recovery trajectories, with socially deprived populations often experiencing poorer health outcomes as a result of associated barriers such as delayed treatment, inadequate rehabilitation resources, and reduced access to support systems (Bhimjiyani et al., 2017; Kristensen et al., 2017; McMaughan et al., 2020). Social-deprivation is also known to influence patient-reported function, pain, anxiety and depression and general mental health in prevalent orthopaedic trauma (Wright et al., 2019; Bhimjiyani et al., 2017) which incurs a large cost to both patients and society and while these disparities are well-documented in broader orthopaedic contexts, their specific influence on femoral PPF recovery remains underexplored.

Since psychosocial factors are culturally mediated, orthopaedic trauma research in femoral PPFs continues to be excessively focused on surgical, clinical and physical components, and there is a growing body of literature that highlights the importance of incorporating patient perspectives into recovery evaluations to give weight of the psychosocial burden associated and thus inform care pathways and improve outcomes more sensitively (Griffiths et al., 2015; Schiller et al., 2015).

### Study Rationale

Given the potential for high complication and mortality risk, poor functional outcome, and high socio-economic burden associated with femoral PPFs, this study aims to address a knowledge gap and contextualise injury and recovery experiences within patients’ broader social worlds to provide deeper understanding of how individuals from varying levels of social deprivation experience recovery following femoral PPF. Identifying barriers to equitable healthcare access, experience and recovery is imperative to provide foundations for targeted and holistic interventions that address systemic disparities and improve patient centred care for all patients who suffer from femoral PPF.

### Aims and Objectives

The primary aim of the study was to explore the injury and recovery experience of patients following femoral PPF, with an emphasis on the influence of social deprivation on this experience to inform targeted holistic care. Specifically, the study’s aims are:

1. Understand the lived experience of femoral PPF injury and recovery before and during the hospital stay with emphasis on psychological and social domains.
2. Investigate how social deprivation influences experiences of femoral PPF injury and recovery with emphasis to healthcare experience before and during hospitalisation.
3. Identify actionable recommendations to improve patient-centred care and inform recovery pathways.

### Outcome

Through addressing these aims and objectives, this study contributes to growing evidence base on equitable and patient-centred care in trauma and orthopaedics. It provides nuanced insights of challenges faced by patients recovering from femoral PPF whilst highlighting the role of recognising socio-economic disparities in shaping outcomes. The study findings aim to inform priorities of healthcare planning in femoral PPF pathways and in guiding interventions for socially deprived populations whilst supporting future research into equitable and holistic recovery approaches.

## Methods

A qualitative study using semi-structured interviews was conducted to explore the experience of recovery following femoral PPF’s. The study was conducted at a single Major Trauma Hospital in the North West of England between August 2022 and August 2023. Participants were recruited and data collection took place on trauma and orthopaedic wards, where participants were admitted for the treatment and care following a femoral PPF. The study utilised Braun and Clarke’s (2006) thematical analysis framework to identify key patterns and themes within the data.

### Study Setting

Semi-structured interviews were conducted in private settings on the trauma and orthopaedic wards, prioritising quiet and private environments to ensure participant confidentiality and comfort. The primary location was the ward’s family room however if this was unavailable or unsuitable due to participant mobility, some interviews were conducted at participant bedsides with closed curtains and with the participant’s consent. Interviews were conducted post-operatively and prior to discharge from hospital, allowing the participants to reflect on their injury and immediate recovery experiences.

### Participant Recruitment

Identification and recruitment of the participants were carried out by the Chief Investigator, who also worked clinically as a physiotherapist on the trauma and orthopaedic wards. Potential participants were identified through routine daily ward handovers, daily accident and emergency admission lists, and daily operating theatre lists. Eligible participants were then approached and provided written and verbal information in regard to the study. Interested participants received written patient information sheets and were provided up to 72 hours to provide written informed consent.

### Eligibility Criteria

- **Inclusion criteria**: Adults aged 18 and over with radiographic evidence of femoral PPF, has full cognitive capacity to consent to involvement in the study, and not under end-of-life care.
- **Exclusion criteria**: Patients outside of the specified age range, or without radiographic evidence of femoral PPF, or lacking cognitive capacity surrounding consenting to their involvement in the study or is receiving end-of-life care.

A purposeful sampling approach was used to recruit participants who could provide detailed insights into their recovery experiences following femoral PPF. Participants were further categorised in socio-economic subgroups based on the English Indices of deprivation, thus enabling comparative analyses across socio-demographic contexts.

### Data Collection

Data was collected via individual semi-structured interviews and were guided by topic guides co-developed with patient and public involvement (PPI) during focus group interviews to ensure relevance and covered key domains including:

1. Experience of the injury and post-operative care.
2. Impact of the injury and subsequent treatment on daily life.
3. Recovery priorities and concerns.
4. Expectations for the future.

Interviews were conducted by the Chief Investigator and audio-recorded using a Dictaphone, and transcribed verbatim. Transcriptions were then anonymised, with identifiable information replaced with pseudonyms to protect participant confidentiality.

### Data Analysis

A thematic analysis approach, informed by Braun and Clarke’s (2006) methodology, was used to analyse the data. Nvivo® software was used to manage, code and develop themes from the transcripts following a multi-step process:

1. **Familiarisation**: Transcripts were first read multiple times to develop a comprehensive understanding of the dataset.
2. **Coding**: Initial coding was informed by line-by-line analysis of narratives to ensure alignment with the study objectives. Deductive coding then took place, involving working down from pre-existing understandings from previous research. Inductive coding was then used to organise significant segments and capture emerging themes.
3. **Theme development**: Themes were then organised into conceptual themes and sub-themes across two categories, with a codebook developed for each socio-economic subgroup. Themes and sub-themes in the two categories were then compared across subgroups to identify similarities, differences and new themes or patterns.

The dataset was analysed repeatedly and meticulously to ensure reliability and consistency. The two categories, namely ‘Recovery Experience’ and ‘Socio-economic Influences’, synthesised four conceptual themes each which captured participants’ recovery and the influence of socio-economic status following femoral PPF.

### Ethical Considerations

The study sought and obtained ethical approval via the Research Ethics Committee, Health Research Authority. All participants provided informed written consent and all research procedures adhered to hospital confidentiality policies alongside the Data Protection Act 2018. Identifiable participant data was secured and stored on Trust premises and networks, and only accessible by members of the authorised research team, whilst regulated by the Caldicott Guardian.

## Results

Braun and Clarke’s thematic analysis framework was utilised in this study to explore patients’ experiences of recovery following femoral periprosthetic fractures and to understand how socio-economic factors may influence these experiences. Four core themes were identified via interviews across two primary categories: “Recovery Experience” and “Socio-economic Influence”. The core themes provide insight into the physical, emotional and social challenges patients may face during recovery from femoral periprosthetic fractures, as well as the role of socio-economic status in shaping these experiences.

### Key abbreviations

- Participant x “HD” – High deprivation participant
- Participant x “MD” – Moderate deprivation participant
- Participant x “LD” – Low deprivation participant

### Category 1: Recovery Experience

#### Theme 1: Persistent Physical and Pain-Related Challenges

##### Definition

This theme portrays the ongoing physical challenges in which the patients face, including pain, mobility or function issues, and the impact on daily life.

##### Quotes and Analysis

Participant 3-HD reported, “The pain was unbearable..”, highlighting the intensity of physical discomfort during the acute phase of the recovery. Similarly, Participant 4-LD shared “I can’t do much right now… I plan to move around with a stick once I feel stronger”, which illustrates how physical limitations restrict activities, thus affecting patients’ daily routines and independence.

##### Interpretation

These accounts emphasize how physical pain and mobility challenges are fundamental to the recovery experience. The need for timely and adequate analgesia and physical rehabilitation is obvious, as patients express a desire to regain former abilities but face setbacks due to ongoing discomfort and dependence.

#### Theme 2: Emotional Impact and Adaptive Resilience

##### Definition

This theme reflects the psychological toll of recovery, including frustration, anxiety and the adaptive resilience required to adapt to new physical limitations.

##### Quotes and Analysis

Participant 1-MD noted, “Mentally, it’s been the hardest… I’ve been feeling down about that being delayed”, reflecting the mental challenges associated with delays of surgery during recovery. Participant 8-MD described feeling “more of a prisoner now”, expressing the psychological strain of dependency on others and the frustration of limited freedom.

##### Interpretation

The emotional struggle patients feel is often associated with the loss of autonomy and reliance on others. Yet, despite these challenges, patients demonstrate adaptive resilience, finding ways to manage their emotional well-being by focusing on gradual improvements and setting recovery goals.

#### Theme 3: Reclaiming independence as a central recovery goal

##### Definition

This theme highlights participants’ aspirations to regain independence, a central motivation driving their recovery efforts.

##### Quotes and Analysis

For Participant 1-MD, “I hope to be back to competing,” captures a strong desire for a return to pre-injury activities. Participant 8-MD also shared, “Being able to get out of the house independently,” indicating a fundamental goal to regain self-sufficiency.

##### Interpretation

Independence emerged as a common aspiration, highlighting the significance of autonomy in participants’ lives. The desire to manage daily tasks, engage in hobbies, and minimise dependence on others emphasises the role of independence in recovery and overall well-being.

#### Theme 4: Healthcare Satisfaction and Desired Improvements

##### Definition

This theme captures participants’ perceptions of healthcare quality, including satisfaction with care and areas needing improvement.

##### Quotes and Analysis

Participant 8-MD described their care as “150% excellent,” demonstrating satisfaction with the support received. However, Participant 4-LD commented, “Doctors should listen to patients more when they say they’re in pain,” highlighting a perceived gap in responsiveness that impacted their experience.

##### Interpretation

Participants generally expressed satisfaction with the care received but noted areas where improvements in communication and attentiveness could enhance the patient experience. This theme suggests that clear communication and patient-centred responsiveness are key components of perceived healthcare quality.

### Category 2: Socio-economic Influences

#### Theme 1: Family Support and Reliance on Social Networks

##### Definition

This theme emphasises the critical role of family and social networks in offering practical and emotional support, particularly for those from higher-deprivation settings.

##### Quotes and Analysis

Participant 2-HD highlighted reliance on family: “My son and daughter-in-law have been helping me”. Likewise, participant 3-HD expressed concern for family safety: “I’m constantly thinking about whether [my wife] is safe”, illustrating the dual dependency on and support for family and friends during recovery.

##### Interpretation

Family and social support played a crucial role in participants’ ability to cope with recovery demands. For participants in high-deprivation contexts, family support appeared especially significant, offsetting limitations in healthcare accessibility.

#### Theme 2: Resource Accessibility and Perceptions of Support

##### Definition

This theme reflects participants’ experiences with accessing healthcare resources, capturing both appreciation and frustration amid socio-economic barriers.

##### Quotes and Analysis

Participant 5-MD noted, “I think I should have had more help at home”, indicating that resources were insufficient to potentially avoid injury. Participant 6-LD stated, “The staff seemed stretched, and there were moments when I felt vulnerable”, highlighting how resource constraints affected patients’ sense of security.

##### Interpretation

Socio-economic constraints restricted participants’ access to resources, affecting their perceptions of support. For most individuals interviewed, resource deficiencies compromised quality of care, especially in more deprived populations, underscoring the necessity for equitable healthcare access in both the community and when recovering from femoral periprosthetic fractures.

#### Theme 3: Healthcare Communication and Coordination Gaps

##### Definition

This theme reflects participants’ experiences with delays, miscommunication, and gaps in healthcare coordination, often linked to socio-economic factors.

##### Quotes and Analysis

“I was supposed to have surgery on Thursday, but it was delayed until Saturday,” explained Participant 3-HD, showing how delayed procedures affected recovery. Participant 1-MD also expressed a preference for quicker communication, sharing, “I would have preferred to be done sooner.”

##### Interpretation

Delays and coordination gaps exacerbated the stress associated with the recovery process, particularly when compounded by socio-economic constraints. This theme highlights the need for improvement in timely communication and patient-centred scheduling to enhance the overall patient experience.

## Discussion

This study examines the recovery of patients after femoral periprosthetic fracture, particularly highlighting the role of socio-economic factors in shaping these experiences. Thematic analysis revealed seven key themes across two categories: “Recovery Experience and “Socio-economic Influences”. This discussion will place these findings within the broader literature, analyse the socio-economic impact on recovery, and provide practical recommendations for enhancing patient-centred care for this demographic.

### Contextualizing Findings within Existing Literature

The study’s findings are consistent with previous studies on recovery following femoral periprosthetic fractures, highlighting significant physical and emotional challenges faced by patients. Consistent with prior research, participants in this study often described persistent physical pain and limited mobility as central obstacles in their recovery process (Demnati et al., 2024; Islam et al., 2022). These physical issues affected not only participants’ functional abilities but also their psychological well-being, with several reporting frustration, anxiety or reduced confidence. For example, Participant 3-HD described their pain as “unbearable”, reflecting the profound toll of recovery on mental resilience. Similarly, Islam et al. (2022) also documented symptoms of increased anxiety and depression among patients recovering from periprosthetic fractures, emphasising the dual physical and psychological dimensions of the recovery process.

This study extends the literature by emphasising the role of independence as a primary recovery goal. Participants, like Participant 8-MD, who prioritised “being able to get out of the house independently”, reflects findings from Beer et al. (2021) and Stott-Eveneshen et al. (2017) who identified independence as central to recovery for patients with orthopaedic injuries. These findings indicate that autonomy serve as a psychological motivator and as a measure of recovery success, thereby supporting recommendations for rehabilitation programs tailored to patient-defined foals (Tobiesen Pedersen et al., 2023)

The significance of family support and social networks in the recovery process emerged as a prominent finding, particularly for participants from socio-economically disadvantaged backgrounds. Participants in high-deprivation settings often relied on family members to manage their recovery needs, aligning with Yujkin et al. (2023), who demonstrated that robust social support networks correlate with improved rehabilitation outcomes. Conversely, participants lacking sufficient family support, the absence of formalised systems presented a significant barrier to recovery.

### Interpreting the Socio-Economic Impact on Recovery

This study underscores the impact of socio-economic status on recovery experiences. Participants from higher-deprivation backgrounds often reported difficulties with healthcare access, resource limitations, and care coordination. These findings align with Kristensen et al. (2017), who found that socio-economic disparities often lead to delayed treatment and unmet needs, adversely influencing recovery outcomes. For instance, Participant 1-MD expressed dissatisfaction with surgical delays, stating, “I would have preferred to be done sooner”. Such delays were further complicated by a perceived lack of communication, intensifying stress and uncertainty during the recovery.

Participants from higher-deprivation groups also noted the significance of responsive and transparent healthcare communication, consistent with McMaughan et al. (2020). For the participants, clear and timely updates from healthcare professionals and providers were essential in mitigating feelings of vulnerability. This suggests that socio-economic barriers heighten emotional strain during recovery, thereby increasing the necessity for patient-centred communication protocols.

### Implications for Healthcare Practice

The findings of this study have several suggestions for healthcare providers and policy makers:

#### 1. Integrated Physical and Psychological Support

Persistent physical and emotional challenges throughout recovery highlight the necessity for comprehensive care models that incorporate mental health and psychological support. Regular evaluations to assess both physical progress and psychological well-being could help improve recovery outcomes (Demnati et al., 2024; Islam et al., 2022). Prioritising tailored and progressive rehabilitation resources, including access to regular physiotherapy, pain management programmes an counselling/psychological services is essential.

#### 2. Patient-Centred Goal-Setting

Setting independence as a recovery goal emphasises the need for patient involvement in personalised goal setting during rehabilitation. Studies such as Beer et al., (2021) and Pedersen et al., (2023) indicates that autonomy-focused methods enhance motivation and improves patient outcomes. Involving patients to participate in defining meaningful benchmarks and goals can increase engagement and promote a sense of control over the recovery process.

#### 3. Strengthening Family and Social Support Mechanisms

The importance of family support highlights the need for formalised systems of support, particularly for those from HD backgrounds. Community based programs, respite care, and caregiver training initiatives could help address gaps in recovery support (Auais et al., 2019; Yukin et al., 2023). For patients lacking family network, providing access to subsidised in-home care services could prove especially beneficial.

#### 4. Improved Communication Protocols

Clear, transparent communication is imperative in building patient trust and satisfaction. Timely updates and responsiveness from healthcare providers is essential, especially during uncertain times such as pre-operatively. Efforts to enhance communication may include designating a single point of contact for patient inquiries, providing clearer timelines for care and recovery in hospital, and proactively addressing issues related to delays (Havana et al., 2023; Sharkiya, 2023).

#### 5. Recognised pathway integration

Currently, femoral PPF fractures are managed jointly by orthopaedic teams alongside either elderly medicine or orthogeriatric specialities which has not yet been standardised nationally. Orthogeriatric or elderly medicine input is aimed to provide more holistic medical intervention for this demographic whilst in acute care however does not constitute against any best practice tariffs attached to the care and coordination of intervention established in patients with neck of femur fractures and therefore can be recognised as a limitation of coordination and treatment in their acute care. Despite national drivers such as the Non Ambulatory Fragility Fracture (NAFF) initiative and framework supporting the involvement of elderly medicine or orthogeriatric teams in this patient population amongst others, efforts are yet to be made to address more strict criteria for timely surgical intervention.

### Study limitations and future research directions

It is important to acknowledge several limitations when interpreting the findings of this study. The qualitative design of the study, while offering comprehensive and detailed insights, restricts the generalisability of its conclusions. It should also be noted that there was only a sole researcher throughout data collection and analysis. Additionally, the small sample size may not adequately capture the diversity of experiences within the population of patients recovering from femoral periprosthetic fractures. Future research efforts should aim to address these limitations by incorporating a larger, more representative sample size, and longitudinal designs to monitor recovery trajectories over time and in the community setting whilst being peer reviewed during the data analysis process.

This study addresses self-reported experiences, which are inherently subjective and may be influenced by individual perceptions. Future research could enhance and compliment these findings with the use of integrated objective measures of recovery outcomes, such as functional assessments or healthcare metrics such as length-of-stay, days awaiting surgery, pain scales, time spent with Allied Health Professional etc. Additionally, research exploring specific interventions such as patient centred communication training for healthcare professionals or socio-economic community support programs would be encouraged in addressing disparities and enhancing recovery outcomes in all orthopaedic injuries.

## Conclusion

This study highlights the complex recovery journey faced by patients with periprosthetic fractures, emphasising the influence of socio-economic factors in their experiences and outcomes. Through qualitative exploration of patient narratives, it is evident that recovery involves not only physical process but is deeply shaped by psychological resilience, family support, and access to resources.

Patients from socio-economically deprived backgrounds faced greater challenges, including limited access to coordinated care and support systems, emphasizing the need for equity-focused healthcare strategies and systems. Regardless of these challenges, striving for independence was a universal recovery goal, further highlighting the importance of patient-centred approaches that focus on individual needs and goals.

This study provides deeper understanding of the recovery process, contributing to the growing body of evidence focused on improving patient-centred care for individuals recovering from periprosthetic fractures. The findings highlight the urgent need for healthcare systems to address disparities and integrate physical, psychological, and social aspects into recovery planning. These insights set the foundation for future initiatives aimed at enhancing the quality and equity of care for this vulnerable population.

## Data Availability

All data produced in the present study are available upon reasonable request to the authors

## DECLERATIONS OF INTERESTS

All authors have completed the ICMJE uniform disclosure form at www.icmje.org/coi_disclosure.pdf and declare that all authors had financial support from the NIHR Applied Research Collaboration North-West Coast internship programme in 2022 for the submitted work.

## DECLERATIONS & ACKNOWELEDGEMENT

Artificial Intelligence-assisted tools were used to support language editing and refinement of the manuscript structure only. All content was reviewed, edited, and approved by the author, who takes full responsibility for the work.

## RESEARCH ETHICS APPROVAL

South Central - Hampshire B Research Ethics Committee of the Health Research Authority (HRA) and Health and Care Research Wales (HCRW) gave ethical approval of this work.

